# Polygenic Scores for Schizophrenia and Educational Attainment Predict Global Functioning Across Psychiatric Hospitalization Among People with Schizophrenia

**DOI:** 10.1101/2025.05.20.25328039

**Authors:** Evan J. Giangrande, Anders Kämpe, Jaana Suvisaari, Markku Lähteenvuo, Emilia Vartiainen, Karita Salo, Olli Pietiläinen, Aarno Palotie, Jordan W. Smoller, Benjamin M. Neale

**Affiliations:** Analytic and Translational Genetics Unit, Massachusetts General Hospital, Boston, MA, USA; Psychiatric and Neurodevelopmental Genetics Unit, Massachusetts General Hospital, Boston, MA, USA; Stanley Center for Psychiatric Research, Broad Institute of MIT and Harvard, Cambridge, MA, USA; Institute for Molecular Medicine Finland, University of Helsinki, Helsinki, Finland; Department of Molecular Medicine and Surgery, Karolinska Institutet, Stockholm, Sweden; Finnish Institute for Health and Welfare, Department of Healthcare and Social Welfare, Helsinki, Finland; Department of Forensic Psychiatry, University of Eastern Finland School of Medicine, Niuvanniemi Hospital, Kuopio, Finland; Neuroscience Center, Helsinki Institute of Life Science, University of Helsinki, Helsinki, Finland

## Abstract

**Key Points:** *Question:* Is variation in global functioning among people with schizophrenia associated with genetic differences?

*Findings:* In this genetic association study of 5991 adults with schizophrenia and 59,795 hospitalizations, polygenic scores for schizophrenia and educational attainment predicted global functioning at psychiatric admission and discharge, as well as the magnitude of functional improvement during hospitalization. Higher polygenic burden for schizophrenia was consistently associated with worse functional outcomes, including less functional improvement during hospitalization, while educational attainment polygenic score showed a more complex pattern of associations.

*Meaning:* Polygenic scores may help disentangle heterogeneity in schizophrenia functioning and course.

**Importance:** Schizophrenia is characterized by heterogeneity in disease outcomes and course. The extent to which this heterogeneity is associated with genetic variation is unclear.

**Objective:** To investigate whether polygenic scores (PGS) for schizophrenia and educational attainment are associated with global functioning during inpatient psychiatric hospitalization among patients with schizophrenia.

**Design, Setting, and Participants:** Adults with schizophrenia were recruited nationwide for the SUPER-Finland Study between 2015 and 2018. Global functioning scores recorded during hospitalizations between 1994 and 2019 were extracted from a complete, longitudinal register. Data for the current genetic association study were analyzed between May 2024 and April 2025. We used linear mixed-effects models to examine associations among PGS and global functioning at admission and discharge, as well as functional change during hospitalization.

**Exposures:** Psychiatric hospitalization and PGS for schizophrenia and educational attainment.

**Main Outcomes and Measures:** Admission global functioning, discharge global functioning, and functional change during hospitalization.

**Results:** We analyzed 117,810 global functioning scores from 59,795 hospitalizations and 5991 participants (2733 [45.62%] female, median [IQR] age = 47 [20] years). Higher schizophrenia PGS predicted lower admission global functioning (**β** = −0.20; 95% CI, −0.40 to 0.00; P = .05) and discharge global functioning (**β** = −0.36; 95% CI,−0.56 to −0.17; P < .001), and less functional improvement (**β** = −0.31; 95% CI, −0.49 to −0.14; P < .001). Higher educational attainment PGS predicted greater functional improvement (**β** = 0.20; 95% CI, 0.03 to 0.37; P = .02) but worse admission global functioning (**β** = −0.23; 95% CI, −0.43 to −0.04; P = .02).

**Conclusions and Relevance:** Higher genetic liability for schizophrenia is associated with worse global functioning across psychiatric hospitalization, including less functional improvement. Integrating PGS and clinically relevant, longitudinal disease outcomes may help parse heterogeneity in schizophrenia prognosis and course.

## Background

People with schizophrenia (SZ) show large individual differences in disease course,^1–4^ symptomatology,^4–7^ global functioning,^3,8–10^ cognitive functioning,^5,11,12^ and treatment response.^13,14^ Schizophrenia is also quite heterogeneous genetically: many variants, including hundreds identified by genome-wide association studies (GWAS), confer risk.^15–18^ Individual genetic risk for developing schizophrenia is often quantified as a polygenic score (PGS), which is a weighted sum of the risk variants a person carries. Compared to PGS for other psychiatric traits, SZ PGS have greater predictive power due to schizophrenia’s relatively high heritability and well-powered discovery GWAS.^15,19,20^

Recently, interest has increased in using PGS to predict not only lifetime schizophrenia risk, but also variation in clinical outcomes among people diagnosed with the disorder.^21–26^ By using PGS to disentangle schizophrenia’s extensive phenotypic heterogeneity, it may be possible to improve clinical stratification, prediction, and prognosis–tasks that will be essential for precision psychiatry but that are difficult using phenotypic indicators alone.^27^ Previous outcomes-focused work has examined associations between SZ PGS and treatment response^28–32^ and resistance,^31,33–35^ psychiatric hospitalization burden,^36–38^ symptom severity,^37,39,40^ diagnostic shifts,^36,39^ and cognitive ability.^12,40–42^ Results have generally indicated that higher polygenic risk for schizophrenia is associated with worse outcomes, although effect sizes and significance have varied across studies. This inconsistency stems from insufficient statistical power–most analyses have relied on sample sizes of only a few hundred cases–and the use of heterogeneous outcome variables, including ad hoc, study-specific measures.^22^ A third limitation is that most past research has focused on antipsychotic treatment response, which, although important, represents only one facet of schizophrenia. More research is needed on other clinically relevant schizophrenia outcomes, ideally using deep, real-world phenotypes.^22,43^ Finally, nearly all prior studies have used cross-sectional phenotypes, which offer only a snapshot of an individual’s disease course and preclude crucial insights about associations of genetic risk and longitudinal change in clinical status. The few previous longitudinal studies have largely focused on first-episode psychosis.

Here, in a sample of 5991 adults with SZ, we tested whether SZ PGS predicted global functioning (GF)–a clinically relevant,^4,9,10,44^ real-world^43^ outcome that has been underexplored in past schizophrenia genetics research–across inpatient psychiatric hospitalizations.

Leveraging complete, longitudinal health record data from a population register^45^ and 117,810 GF measures from 59,795 hospitalizations, we analyzed the extent to which SZ PGS predicted GF at psychiatric admission and discharge, as well as individual change in functioning during hospitalization. In addition to SZ PGS, we also analyzed PGS for educational attainment (EA), a trait with extremely well-powered PGS^46^ and strong, primarily negative, phenotypic and genetic associations with SZ.^47,48^ By applying robust statistical models to an unprecedentedly large number of repeated clinical outcome measures, we were able to parse schizophrenia heterogeneity in global functioning and response to hospitalization while addressing limitations of previous outcomes-focused PGS studies.

## Methods

This study followed Strengthening the Reporting of Genetic Association Studies (STREGA)^49^ guidelines.

### Sample

We analyzed data from the SUPER-Finland study.^50^ From November 2015 to December 2018, 10,417 adults with a history of at least one psychotic episode were recruited nationwide by psychiatrists and psychiatric nurses at inpatient and outpatient psychiatric facilities as well as primary care clinics and newspaper advertisements. Participants provided written consent, and the Ethics Committee of the Hospital District of Helsinki and Uusimaa approved the study. The current study focused on a subset of 5991 individuals with a history of schizophrenia or schizoaffective disorder diagnosis, available global functioning scores, and PGS data (eFigure 1). Diagnoses were verified using *International Statistical Classification of Diseases and Related Polygenic Prediction of Global Functioning in Schizophrenia*.

**Figure 1.**
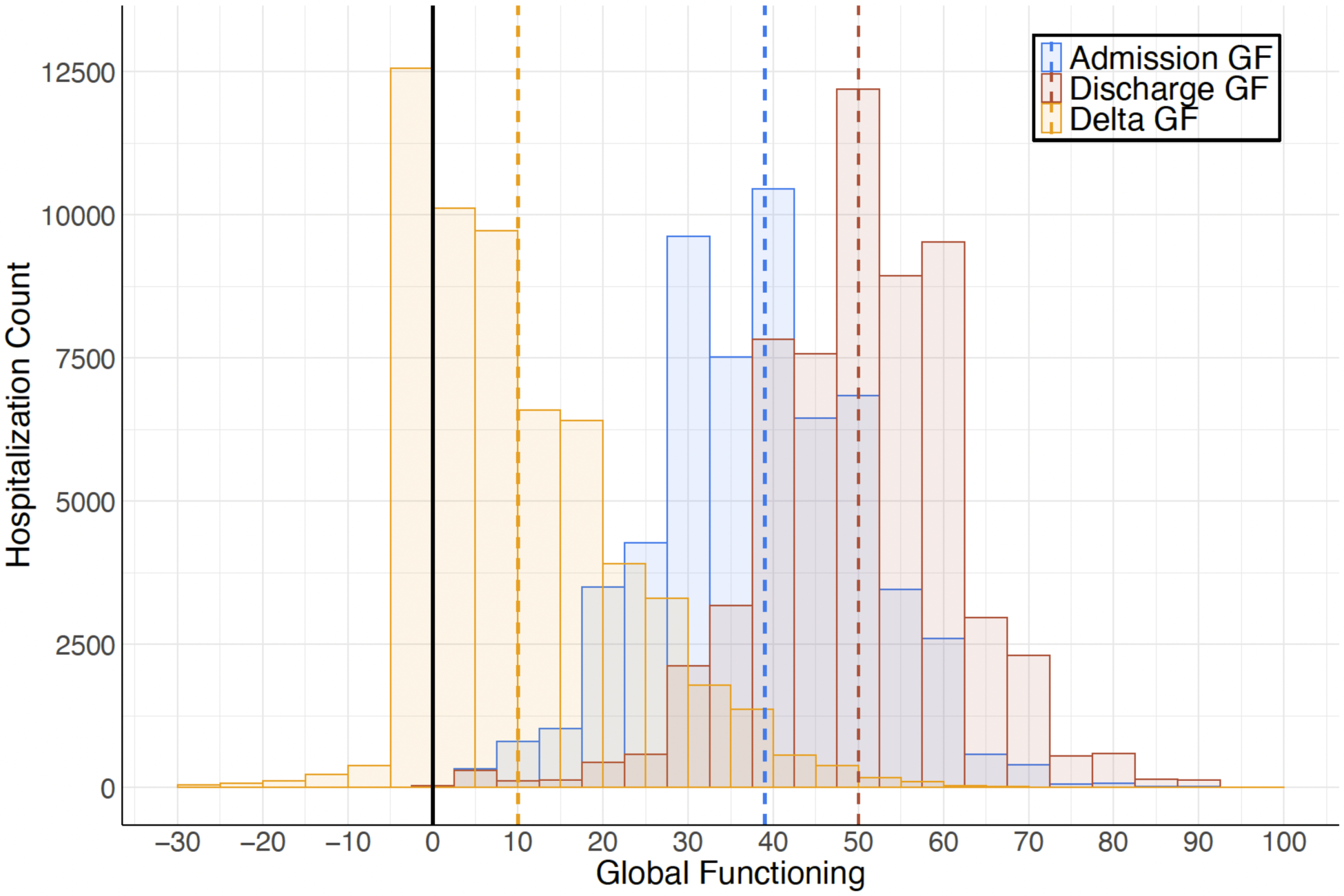
Global Functioning Across 59,795 Psychiatric Hospitalizations. GF: global functioning. Delta GF = Discharge GF - Admission GF. Dashed vertical lines depict median GF scores.

*Health Problems* (ICD) codes^51–53^ from the Care Register for Health Care, HILMO, which includes data from every hospitalization or visit to a specialty outpatient clinic in Finland since 1969 and 1998, respectively, and the Register of Primary Health Care Visits, which includes all nationwide primary care visits since 2011^54^ (eMethods). Participants had at least one lifetime, inpatient psychiatric hospitalization as documented in HILMO. These psychiatric hospitalization records are complete–we can assume that no hospitalizations are missing–and longitudinal, enabling us to capture full hospitalization histories for each participant.^54^

### Phenotype Measures

#### Global Functioning

GF is a transdiagnostic estimate of how well or poorly a patient is doing in their day-to-day lives including their mental health status, symptom severity, social functioning, and self-care ability. SZ is associated with GF impairment, but people with SZ show substantial heterogeneity in GF.^9,10^ In Finland, GF has been assessed routinely at inpatient psychiatric admission and discharge since 1994 using the Global Assessment Scale–a clinician-rated, 1-to-100 scale for which higher scores indicate better functioning^55,56^ (eMethods). From HILMO, we extracted adult psychiatric admission and discharge GF scores for each SUPER-Finland participant from 59,795 hospitalizations (117,810 total admission and discharge GF scores) between January 1994 and February 2019 (Figure 1). To estimate functional change during a given hospitalization while accounting for baseline differences in functioning, we also analyzed Discharge GF adjusted for Admission GF (Change GF). Missingness was low across the GF variables and can be considered random (eMethods).

#### Deep Phenotypes

To explore the GF scores’ clinical relevance in our sample, we also examined their associations with a variety of important phenotypic indicators (Figure 2; eMethods; eTable 1). From HILMO, we extracted median length of psychiatric stay as well as number of psychiatric hospitalizations and number of days in the psychiatric hospital in the five years following a participant’s first psychiatric hospitalization. Educational attainment, difficulties in school, and suicide attempt history were self-reported at study entry (eMethods). Two subtests of the Cambridge Neuropsychological Test Automated Battery (CANTAB) were administered at study entry (eMethods): Paired Associates Learning, which measures visual memory, and Reaction Time.^57–59^

**Figure 2.**
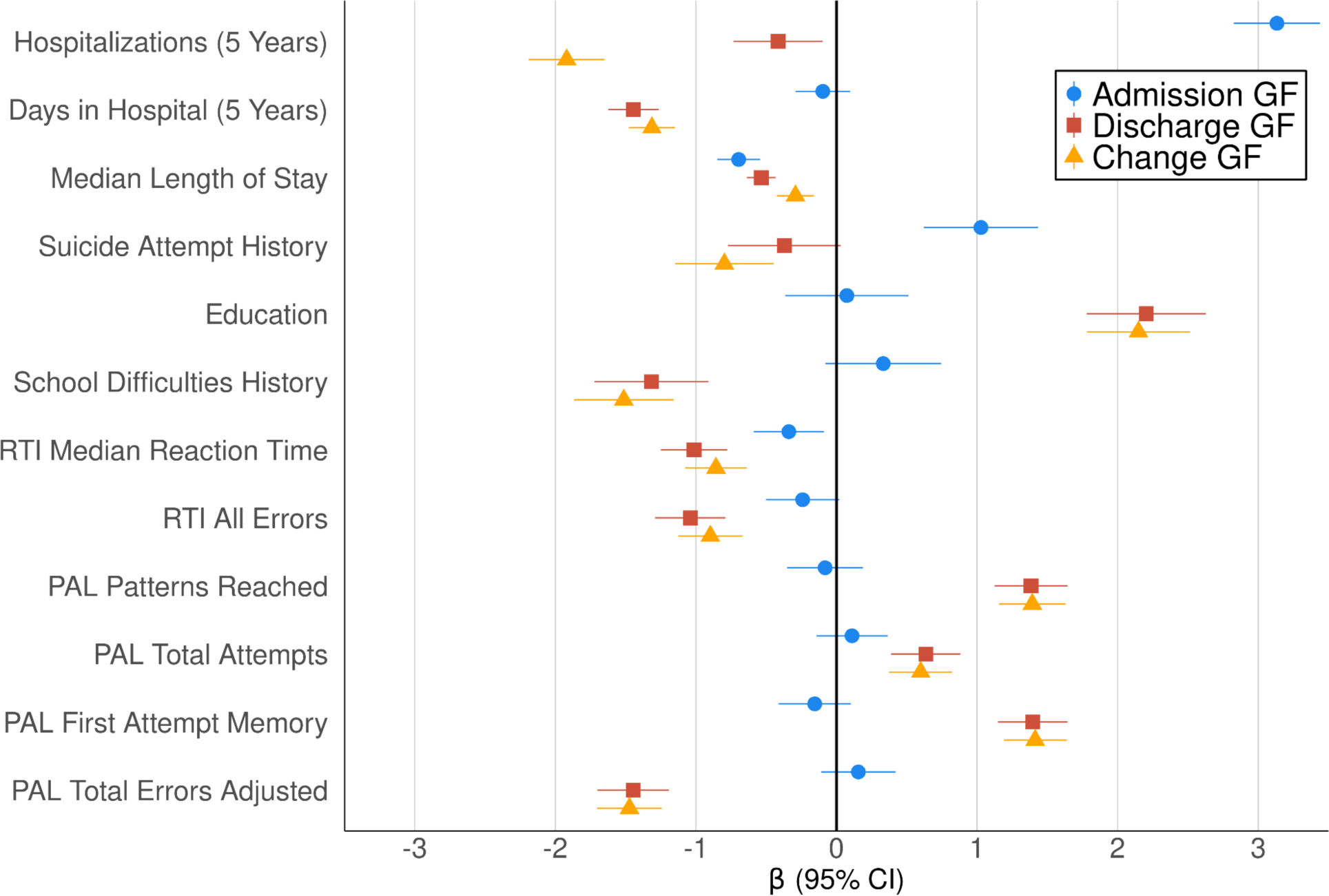
Phenotypic Associations Among Global Functioning Scores and Deep Phenotypes. GF: global functioning. Change GF = Discharge GF adjusted for Admission GF. RTI: Reaction Time. PAL: Paired Associates Learning. 5 Years: these variables were calculated for psychiatric hospitalizations within the initial 5 years following a participant’s first admission (Supplement). RTI and PAL were assessed at study entry (between November 2015 and December 2018). Bars depict 95% CIs.

### Genetic Data

Genotyping was performed at the Broad Institute of MIT and Harvard using the Illumina Global Screening Array (version 1). Quality control and imputation details are presented in the eMethods. The analysis was limited to individuals of European ancestry due to the small number of non-European ancestry participants (n = 71). GWAS summary statistics from Trubetskoy et al. (2021)^15^ and Okbay et al. (2022)^46^ were used to construct PGS for SZ and EA, respectively. SUPER-Finland participants were not part of either GWAS. We chose to analyze EA PGS because it is extremely well-powered among behavioral and psychiatric PGS^46^; EA can be measured robustly at scale and is often reduced among people with SZ^48^; and EA has strong, primarily negative genetic associations with SZ.^47^ We restricted PGS construction to SNPs with a minor allele frequency of ≥ 1% and an imputation score of ≥ 0.95. We used MegaPRS,^60^ which performs well for psychiatric PGS construction,^61^ following the developers’ recommended settings (BLD-LDAK heritability model, BayesR-SS for prediction model construction). Pseudo cross-validation was used to select suitable model parameters, and the 1000 Genomes non-Finnish European population group was used to estimate SNP-SNP correlations.

### Statistical Analysis

#### Linear Mixed-Effects Models

We estimated the influence of PGS on global functioning during hospitalization by fitting separate linear mixed-effects models to Admission GF, Discharge GF, and functional change (Change GF) scores using the nlme R package^62,63^ (eMethods). Both SZ and EA PGS were included as fixed effects in each model, allowing us to estimate the independent association of each PGS. To leverage the repeated GF observations, hospitalizations were nested within individual participants using a random intercept. We accounted for autocorrelation in the residuals by specifying a first-order residual autocorrelation structure in continuous time, which, unlike discrete-time solutions, handled variability in the temporal spacing of hospitalizations.^64^ All analyses employed restricted maximum likelihood estimation (REML) and included the following covariates: sex, year of birth (to control for possible differences across generational cohorts), and the first 10 genetic principal components. Analyses of Discharge GF and functional change also controlled for length of hospitalization stay. To correct for multiple comparisons across our six primary comparisons (2 PGS predictors x 3 GF outcomes), we applied the Benjamini-Hochberg procedure^65^ to control the false-discovery rate (FDR) at .05.

#### Sensitivity Analysis

To examine whether PGS associations were consistent across GF strata, we performed an exploratory, post-hoc sensitivity analysis in which we subset hospitalizations into Admission GF tertiles and re-ran our genetically informed models separately within each tertile (eMethods).

## Results Demographic and Descriptive Information

We analyzed data from 5991 participants including 2733 (45.62%) female participants and 3258 (54.38%) male participants (eMethods; eTable 1). Median age at the beginning of the study was 47 years (IQR = 20). Participants had a median of 6 hospitalizations (IQR = 9) for which GF data was available. Median length of psychiatric stay was 14 days (IQR = 38).

### Global Functioning Across Psychiatric Hospitalization

Median GF score at admission was 39 (IQR = 16), corresponding to major impairment in several areas or some impairment in reality testing or communication (Figure 1).^55,56^ Median Discharge GF score was 50 (IQR = 16), indicating serious symptoms or impairment.^55,56^ Admission GF showed significantly greater variance than Discharge GF (SD = 12.31 and 11.85, respectively; *F*(1, 117,808) = 378.97, *P* < .001; eMethods). Psychiatric hospitalization generally resulted in substantial functional improvement: GF increased during hospitalization by a median of 10 (IQR = 19). GF improved between admission and discharge in 44,539 (76.77%) of hospitalizations (Figure 1). 12,266 (21.14%) of hospitalizations resulted in no improvement, and GF decreased between admission and discharge in 1210 (2.09%) of hospitalizations.

### Global Functioning Associated with Important Phenotypic Outcomes

GF was associated with a variety of other clinically relevant phenotypic indicators (Figure 2). Participants who showed greater Discharge GF and functional improvement (Change GF) had fewer psychiatric hospitalizations and days in the hospital in the 5 years following their first psychiatric admission, shorter median length of stay, less suicidality, higher EA and less likelihood of school difficulties, and better performance on Reaction Time and Paired Associates Learning. Admission GF exhibited weaker associations, likely due to its larger variance than Discharge GF and functional change (eMethods). Higher Admission GF was associated with more psychiatric hospitalizations over the 5 years following one’s first admission but shorter overall length of stay, more suicidality, and quicker reaction time.

due to the inclusion of Admission GF as a covariate.

### SZ and EA PGS Predicted Global Functioning Across Hospitalization

Results from our linear-mixed effects models indicated that higher polygenic risk for schizophrenia predicted significantly worse GF at psychiatric admission (**β** = −0.20; 95% CI, −0.40 to 0.00; P = .05) and discharge (**β** = −0.36; 95% CI,−0.56 to −0.17; P < .001) (Figure 3; eTables 2-5), although the Admission GF association did not survive correction for multiple comparisons (adjusted P = .06). Higher SZ PGS was also associated with less functional improvement during hospitalization (**β** = −0.31; 95% CI, −0.49 to −0.14; P < .001). Higher EA PGS predicted significantly worse functioning at psychiatric admission (**β** = −0.23; 95% CI, −0.43 to −0.04; P = .02) but greater functional improvement (**β** = 0.20; 95% CI, 0.03 to 0.37; P = .02). The association of EA PGS and Discharge GF was positive, but not significant (**β** = 0.15; 95% CI, −0.04 to 0.35; P = .12). Fixed effects including PGS collectively explained 2% of the variance in both Admission GF and Discharge GF (marginal pseudo-R2 = 0.02; eMethods). Marginal pseudo-R2 was higher (0.43) for the functional change analysis The sensitivity analysis indicated that PGS associations were generally consistent across GF tertiles (eMethods; eFigure 2; eTables 6-8). SZ PGS was significantly negatively associated with Discharge GF and Change GF across tertiles. EA PGS associations with Discharge GF and Change GF were positive across tertiles but not significant, likely due to the power decrease from analyzing only one third of the total hospitalizations. Admission GF associations were also attenuated, which may reflect reduced power and decreased outcome variability due to conditioning the analysis on Admission GF tertile.

**Figure 3.**
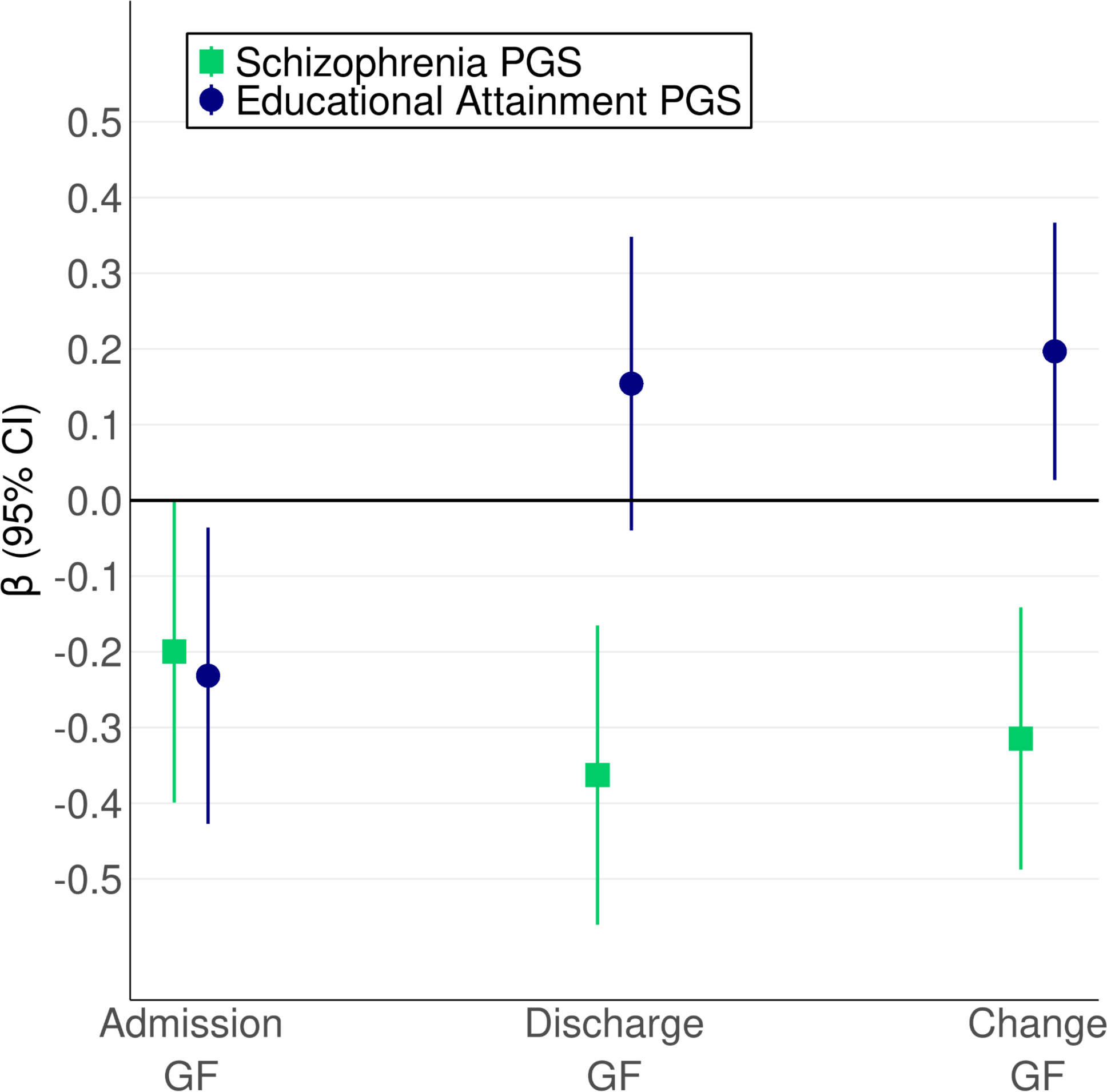
Schizophrenia and EA PGS Predict Global Functioning Across Hospitalization. PGS: polygenic score. GF: global functioning. Change GF: Discharge GF adjusted for Admission GF. Bars depict 95% CIs.

## Discussion

Schizophrenia’s extensive heterogeneity in prognosis and course is widely acknowledged, but its causes are poorly understood. This ambiguity impedes clinical practice and the implementation of research findings. Previous studies have attempted to address these challenges by examining the influence of polygenic scores on schizophrenia clinical outcomes, but most have been limited by small sample sizes and inconsistent, cross-sectional phenotyping. Leveraging complete (i.e., no missingness), longitudinal records from 59,795 psychiatric hospitalizations; 117,810 repeated clinical outcome measures, and a deeply phenotyped sample, this study investigated whether polygenic scores for schizophrenia and educational attainment predicted global functioning across inpatient psychiatric hospitalization among adults with schizophrenia. Phenotypically, GF scores were associated with important variables including hospitalization burden, educational attainment and school difficulties, and cognitive functioning. Higher SZ PGS was consistently associated with lower functioning: SZ PGS predicted lower GF at admission and discharge and less functional improvement during psychiatric hospitalization. EA PGS exhibited a more complicated pattern of associations, with higher EA PGS predicting better functioning at discharge (not significant) and greater hospitalization-related improvement in GF but lower GF on admission. Overall, this study demonstrated the utility of incorporating deeper, clinically relevant outcomes and genetic data to help parse heterogeneity in schizophrenia course and prognosis. Our findings provide new insights into genetic influences on global functioning in schizophrenia, a real-world outcome of key interest to clinicians, researchers, and patients that has been understudied in prior genetically informed research.

Our SZ PGS results are broadly consistent with evidence from previous outcomes-focused studies–particularly adequately powered studies, of which there have been few–that higher SZ PGS is associated with worse schizophrenia outcomes.^22^ Thus, genetic factors that increase lifetime risk for developing schizophrenia also appear to be associated with a more severe clinical presentation and disease course. In contrast to SZ PGS associations, higher EA was associated with better hospitalization response and discharge functioning. Somewhat surprisingly, higher EA PGS predicted worse functioning at admission. Genetically informed studies of EA in the context of schizophrenia have yielded counterintuitive findings^66^ and our results may reflect this complexity. Individuals with higher EA PGS may function relatively well when stable but then decline sharply, or may avoid hospitalization until their functioning is severely impaired. Either scenario would generate a negative association with Admission GF, but we could not test this because we lacked data on GF outside hospitalization.

When deciding whether to refer or admit a patient with schizophrenia for psychiatric hospitalization, clinicians often consider the extent to which the patient would benefit from admission. Results from our novel longitudinal analyses of functional change in response to hospitalization suggest that genetics may be relevant to that decision-making process: differences in hospitalization-related functional improvement among patients with schizophrenia were associated with genetic differences. These findings represent an exciting expansion of the genetically informed literature on schizophrenia treatment response, which has focused primarily on antipsychotic response, to include hospitalization outcomes. More generally, our functional change findings demonstrate the value of analyzing longitudinal change in schizophrenia presentation, a fundamental element of disease heterogeneity that has rarely been explored using genetically informed designs.^21–24^

Focusing on prognostic, rather than diagnostic, genetic prediction enabled us to dissect schizophrenia heterogeneity that is untapped by traditional case-control designs. Collectively, our results suggest that integrating well-powered genetic findings with real-world, clinically relevant outcome phenotypes may help stratify people with schizophrenia who present for psychiatric admission. Outcomes-focused genetics research of this sort will be critical as the field strives toward precision psychiatry and the clinical implementation of psychiatric genetics. Fortunately, it is becoming increasingly feasible to perform outcomes-focused genetic analyses at scale as longitudinal health record data are combined with genomic data.^27^

This study had several strengths. First, our analysis focused on real-world global functioning, a widely-used outcome directly relevant to schizophrenia treatment, phenomenology, and nosology. We analyzed GF scores collected systematically as a standard part of Finnish psychiatric practice. Second, we performed novel longitudinal analyses that captured genetic influences on hospitalization-related functional change using robust statistical models and complete psychiatric hospitalization data. Third, our sample (N = 5991) was considerably larger than most previous outcomes-focused PGS studies of schizophrenia. Our multilevel modeling approach further boosted statistical power by pooling 117,810 repeated GF scores from 59,795 psychiatric hospitalizations. Fourth, the SUPER-Finland sample was deeply phenotyped, enabling us to explore the GF scores’ clinical correlates. Finally, we focused on SZ and EA PGS, two of the best-powered PGS for behavioral traits.

This analysis was limited to a Northern European sample of exclusively European ancestry. Our findings therefore await replication in populations from other ancestries, demographic backgrounds, and geographical locations. A second limitation is that we analyzed participants with a hospitalization history, and GF scores were collected during psychiatric admissions (i.e., the most severe illness periods). The extent to which our findings generalize across the full schizophrenia severity spectrum is therefore unclear. Third, global functioning is a broad index of clinical status and our results therefore do not elucidate specific clinical factors driving observed associations with PGS. Finally, consistent with other psychiatric genetics findings, the genetic associations we observed were small, indicating that PGS are not sufficiently predictive to warrant their integration into schizophrenia care at this time.^20,26^ It is unlikely that currently powered SZ or EA PGS alone would substantially improve prediction of functional status compared to non-genetic indicators.^37^ PGS predictiveness may improve as genomic sample sizes increase and genetic data are integrated with deeper phenotypes^20,67,68^ and it will eventually be possible to calculate well-powered, outcome-specific PGS, but the clinical implementation of genetic results remains premature.

### Conclusions

This study’s findings suggest that higher polygenic liability for schizophrenia is associated with worse functioning at psychiatric admission and discharge, as well as less functional improvement in response to hospitalization. Genetic factors associated with higher educational attainment were associated with worse admission functioning, but better functioning at discharge. Integrating PGS with clinically relevant schizophrenia outcomes helps disentangle heterogeneity in disease severity and course. Further outcomes-focused genetics research may hold promise for improving genetic discovery and clinical decision-making, but PGS predictiveness is currently insufficient for clinical implementation.

## Data Availability

Deidentified data from the SUPER-Finland cohort is available upon reasonable request to Drs. Palotie and Pietiläinen. Data for SUPER-Finland participants who provided consent for biobank participation are available from the THL Biobank (https://thl.fi/en/research-and-development/thl-biobank/for-researchers/sample-collections/super-study). Researchers may apply for access to register data from the Finnish Social and Health Data Permit Authority (Findata; https://findata.fi/en/). Code used to perform analyses and generate figures is available at https://github.com/egiangrande/SUPER_GlobalFunctioning_SZ.

## Acknowledgments

Dr. Giangrande had full access to all the data in the study and takes responsibility for the integrity of the data and the accuracy of the data analysis. We would like to thank the SUPER-Finland Study participants and the SUPER-Finland study nurses and staff. We acknowledge those involved with collecting and maintaining the Finnish health register data, particularly the Care Register for Health Care (HILMO) and the Register of Primary Health Care Visits, and the clinicians who completed the real-world global functioning assessments. We thank the Broad Institute of MIT and Harvard Genomics Platform for conducting genotyping, and the Stanley Center for Psychiatric research for supporting both SUPER-Finland and the present study.

## SUPER-Finland Researchers

The following individuals were instrumental in the execution of the SUPER-Finland study: Aija Kyttälä, Annamari Tuulio-Henriksson, Ari Ahola-Olli, Asko Wegelius, Auli Toivola, Erkki Isometsä, Huei-yi Shen, Imre Västrik, Jari Tiihonen, Jarmo Hietala, Jouko Lönnqvist, Juha Veijola, Jussi Niemi-Pynttäri, Kaisla Lahdensuo, Katja Häkkinen, Kimmo Suokas, Mark Daly, Minna Holm, Noora Ristiluoma, Olli Kampman, Risto Kajanne, Steven E. Hyman, Tarjinder Singh, Teemu Männynsalo, Tiina Paunio, Tuomas Jukuri, Tuula Kieseppä & Willehard Haaki

