## Supplementary Material for "Polygenic Scores for Schizophrenia and Educational Attainment Predict Global Functioning Across Psychiatric Hospitalization Among People with Schizophrenia"

**eMethods**

-Diagnostic Procedure (p.2)
-Global Functioning (p.2)
-Global Functioning Variance Comparison (p.2)
-Deep Phenotypes (p.2)
-Phenotypic Associations (p.3)
-Genotype Quality Control (p.3)
-Linear Mixed-Effects Models (p.3)
-Autocorrelation Parameter Estimates (p.4)
-Sensitivity Analysis (p.4)
-Pseudo-$R^2$ Calculation and Results (p.4

**eFigures**

-eFigure 1. SUPER-Finland Sample Flow Diagram (p.5)
-eFigure 2. PGS Associations Stratified by Admission GF Tertile (p.6)

**eTables**

-eTable 1. Demographic and Descriptive Information (p.7)
-eTable 2. Estimates from Admission GF Linear Mixed-Effects Model (p.8)
-eTable 3. Estimates from Discharge GF Linear Mixed-Effects Model (p.9)
-eTable 4. Estimates from Change GF Linear Mixed-Effects Model (p.10)
-eTable 5. Multiple Comparisons Correction (p.11)
-eTable 6. Estimates from Tertile-Stratified Admission GF Linear Mixed-Effects Model (p.12)
-eTable 7. Estimates from Tertile-Stratified Discharge GF Linear Mixed-Effects Model (p.15)
-eTable 8. Estimates from Tertile-Stratified Change GF Linear Mixed-Effects Model (p.18)

**eReferences**

### eMethods

#### Diagnostic Procedure

Diagnoses were based on ICD codes extracted from the Care Register for Health Care (HILMO) and the Register of Primary Health Care Visits. This approach yields highly accurate schizophrenia diagnoses.[1] Following the diagnostic procedure used in the broader SUPER-Finland study,[2] lifetime diagnoses were assigned according to a hierarchical framework in which psychotic disorders range from major depressive disorder with psychosis, to bipolar disorder, to schizoaffective disorder, to schizophrenia. Schizophrenia is at the bottom of the hierarchy. For individuals with multiple lifetime diagnoses, their final diagnosis is the lowest-hierarchy diagnosis, with schizophrenia trumping all other diagnoses. For example, individuals with a history of bipolar disorder and schizophrenia were designated as having schizophrenia. Consistent with leading genomic consortia,[3] individuals with both schizophrenia and schizoaffective disorder were included.

#### Global Functioning

Since 1994, global functioning has been collected as a routine part of psychiatric inpatient admission and discharge in Finland.[4] Global Assessment Scale (GAS) scores[5] are assigned by clinicians who are all held to the same standards of practice and who undergo similar psychiatric training. The same GAS rating guidelines are followed nationwide.[4] GAS scores range from 1 to 100, with higher scores indicating better functioning. The GAS is analogous to other widely used functional measures including the Global Assessment of Functioning (GAF). Its reliability is acceptable,[5] including in clinical settings where GAS scores are collected longitudinally by multiple raters,[6] and sufficient for analyses of group-level change in functioning over time.[7]

Because GF scores were not collected systematically until January 1, 1994, we excluded a small number of sporadic GF scores collected prior to that date. We observed low rates of missingness in the remaining scores, which can be considered missing at random. 7.37% and 5.03% of post-1993 psychiatric admission and discharge GF scores were missing, respectively. The discrepancy in missingness rates (i.e., higher rate of Admission GF missingness) was driven by hospitalizations where admission occurred in 1993 (immediately prior to the start of systematic GF collection) and discharge took place after the turn of the new year.

In situations where hospitalizations were immediately adjacent (e.g., discharge followed by admission on the same day), we merged hospitalizations and extracted the first admission GF score and the last discharge GF score. In addition to the 1-100 score range, the GAS allows clinicians to assign a zero if they did not have enough information to assess a patient's global functioning. We recoded zeroes to NA prior to analysis.

For SUPER-Finland participants who met our inclusion criteria, a total of 117,810 admission and discharge GAS scores were available. The most recent score available was collected in February 2019.

#### Global Functioning Variance Comparison

To compare the variances of Admission GF and Discharge GF (SD = 12.31 and 11.85, respectively), we performed a Brown-Forsythe test.[8] The test was significant ($F(1, 117{,}808) = 378.97$, $P < .001$), indicating that phenotypic variance in Admission GF was significantly larger than Discharge GF variance. This result suggests that variance decreased as patients stabilized and clinicians gathered additional information about their functional status

#### Deep Phenotypes

We examined associations between GF scores and a variety of phenotypes available in the SUPER-Finland study, prioritizing clinically relevant outcomes.

From the Care Register for Health Care, HILMO, we calculated psychiatric hospitalization burden including number of hospitalizations, total days in the hospital, and median length of stay. We focused on psychiatric hospitalizations during the five years following participants' first hospitalization because nearly all the sample had at least five years of follow-up data available.

SUPER-Finland participants completed an interview that was based on questions from the Finnish Health 2000 and 2011 general population surveys.[9,10] We analyzed educational attainment from the interview and in a small number of cases where it was unavailable, it was extracted from the Register for Completed Education and Degrees. Following the population surveys, educational attainment was coded as one of three levels: basic (compulsory education; no completion of vocational school or the matriculation examination), secondary (passed matriculation examination but no degree; completion of vocational school), and higher (degree from university, polytechnic institute, or higher vocational institution). History of difficulties at school that lasted for more than a year was also collected during the interview. Suicide attempt history was collected using a questionnaire.

Cognitive ability was assessed at entry to the SUPER-Finland study (i.e., between November 2015 and December 2018) using two, Finnish-adapted subtests of the Cambridge Neuropsychological Test Automated Battery (CANTAB): Paired Associates Learning (PAL), which measures visual memory, and Reaction Time (RTI).[11] We analyzed only CANTAB data that were deemed "Complete and Reliable" by research supervisors. For the RTI, which was the five-choice version, better performance is indicated by quicker median reaction time and fewer errors. Better performance on the PAL is indicated by more patterns reached, more total attempts, higher first attempt memory, and fewer total errors. We used an adjusted total errors metric that estimated the number of errors a participant would have made had they completed all trials.

**Phenotypic Associations**
To examine the phenotypic correlates of our GF scores, we performed an exploratory analysis investigating their associations with the deep phenotypes listed above. Specifically, we fit a series of linear mixed-effects models in which each deep phenotype was modeled as a predictor of each GF score. Analogously to our genetic analyses, hospitalizations were nested within individual participants using a random intercept. All analyses controlled for sex and year of birth.

**Genotype Quality Control and Imputation**
Samples were genotyped in four batches at the Broad Institute of MIT and Harvard using the Illumina Global Screening Array (version 1). Within-batch QC excluded samples with poor genotype quality, genetically inferred sex mismatching self-reported sex, sample missingness > 0.05, and Hardy-Weinberg equilibrium (HWE) $P < 1 \times 10^{-12}$. Batches were then merged before undergoing another round of QC, which applied the same filters except the HWE and missingness thresholds were set at $P < 1 \times 10^{-9}$ and > 0.02, respectively. We excluded variants with minor allele frequency (MAF) < 0.01. PLINK 2[12] was used to calculate genetic principal components. Seventy-one participants with non-European ancestry as inferred from the principal components were excluded, as this sample size was insufficient for robust genetic analyses. Imputation was performed using Sequencing Initiative Suomi (SiSu) v.3, a Finnish-specific reference panel.[13] Variants with INFO score < 0.8 were excluded.

**Linear Mixed-Effects Models**
We fit a series of linear mixed-effects models using the nlme R package.[14,15] Separate models were fit to global functioning scores at psychiatric admission and discharge, as well as functional change in response to hospitalization (Change GF; Discharge GF adjusted for Admission GF). For hospitalization $i$ and individual $j$:

$$\mathrm{GF}_{ij} = \beta_0 + \beta_1\mathrm{SZ}_j + \beta_2\mathrm{EA}_j + \beta_3\mathrm{Sex}_j + \beta_4\mathrm{YOB}_j + \mathbf{X}_j\mathbf{b}_X + u_{0j} + \varepsilon_{ij} \quad (1)$$

We were primarily interested in the fixed effects of schizophrenia (SZ) and educational attainment (EA) polygenic scores. Sex (coded as Female/Male), year of birth (YOB), and the first 10 genetic principal components are included as fixed covariates. $X$ is a design matrix of the principal components and b is a vector containing their regression coefficients. Discharge GF and functional change analyses also controlled for the log of length of stay at hospitalization $i$ (not shown), and the analysis of functional change in response to hospitalization controlled for Admission GF (not shown). To account for the repeated GF measures, hospitalization is nested within participant j using random intercept $u0_j$. Autocorrelation in residuals $\epsilon_{ij}$ of GF across hospitalizations is handled by specifying a first-order autocorrelation structure in continuous time, which is robust to unequal temporal spacing of observations.[16] The correlation between residuals at any pair of hospitalizations $i$ and $k$ for individual $j$ is modeled as:

$$\mathrm{Cor}(\varepsilon_{ij}, \varepsilon_{kj}) = \rho^{|t_{ij} - t_{kj}|} \quad (2)$$

Autocorrelation parameter $\rho$ decays exponentially with the time elapsed between discharge dates $t_{ij}$ and $t_{kj}$ such that residual correlations are stronger among temporally closer GF measures and weaker among more distal hospitalizations.

**Autocorrelation Parameter Estimates**

Results from our linear mixed-effects model analyses are discussed in the main manuscript. Parameter estimates are presented in eTables 2-4.

Autocorrelation parameter estimates ($\rho$) equaled 0.93, 0.96, and 0.93 in the Admission GF, Discharge GF, and Change GF analyses, respectively. These estimates indicate strong positive autocorrelation in the residuals of the repeated GF measures, which we modeled as a first-order residual autocorrelation structure in continuous time.

**Sensitivity Analysis**

To confirm that PGS associations were consistent across GF strata, we performed an exploratory, post-hoc sensitivity analysis in which we separated hospitalizations based on Admission GF tertile and re-ran our linear mixed-effects models within each tertile. Lower, Middle, and Upper tertiles corresponded to Admission GF scores ≤ 32, between 33 and 45 (inclusive), and ≥ 46, respectively. Results are presented in eFigure 2 and eTables 6-8. SZ PGS associations with Discharge GF and functional change were significantly negative across all tertiles ($P$s < .05), consistent with results from our main analysis. As in the main analysis, EA PGS was positively associated with Discharge GF and functional change across all tertiles. However, none of these associations were significant ($P$s > .05), unlike the main analysis in which the functional change association was significant. This attenuation of significance is likely driven by power reductions due to analyzing only one third of available hospitalizations. No Admission GF associations were significant ($P$s > .05), reflecting the power reduction and the decreased Admission GF variability as a result of conditioning the sensitivity analysis on Admission GF tertile.

**Pseudo-$R^2$ Calculation and Results**

To estimate the proportion of variance explained by our models, we calculated pseudo-$R^2$ values[17] and the MuMIn package.[18] This approach yields two estimates: marginal $R^2$, which estimates the proportion of variation in GF explained by the fixed effects alone, and conditional $R^2$, which estimates the proportion of variance explained by both fixed and random effects. Conditional $R^2$ for the Admission GF, Discharge GF, and functional change analyses equaled 0.02, 0.02, and 0.24, respectively. Marginal $R^2$ equaled 0.27, 0.26, and 0.43, respectively (eTables 2-4). Our observation of larger pseudo-$R^2$ values for the functional change analysis was driven by the inclusion of Admission GF in the model.

#### eFigures

##### eFigure 1. SUPER-Finland Sample Flow Diagram

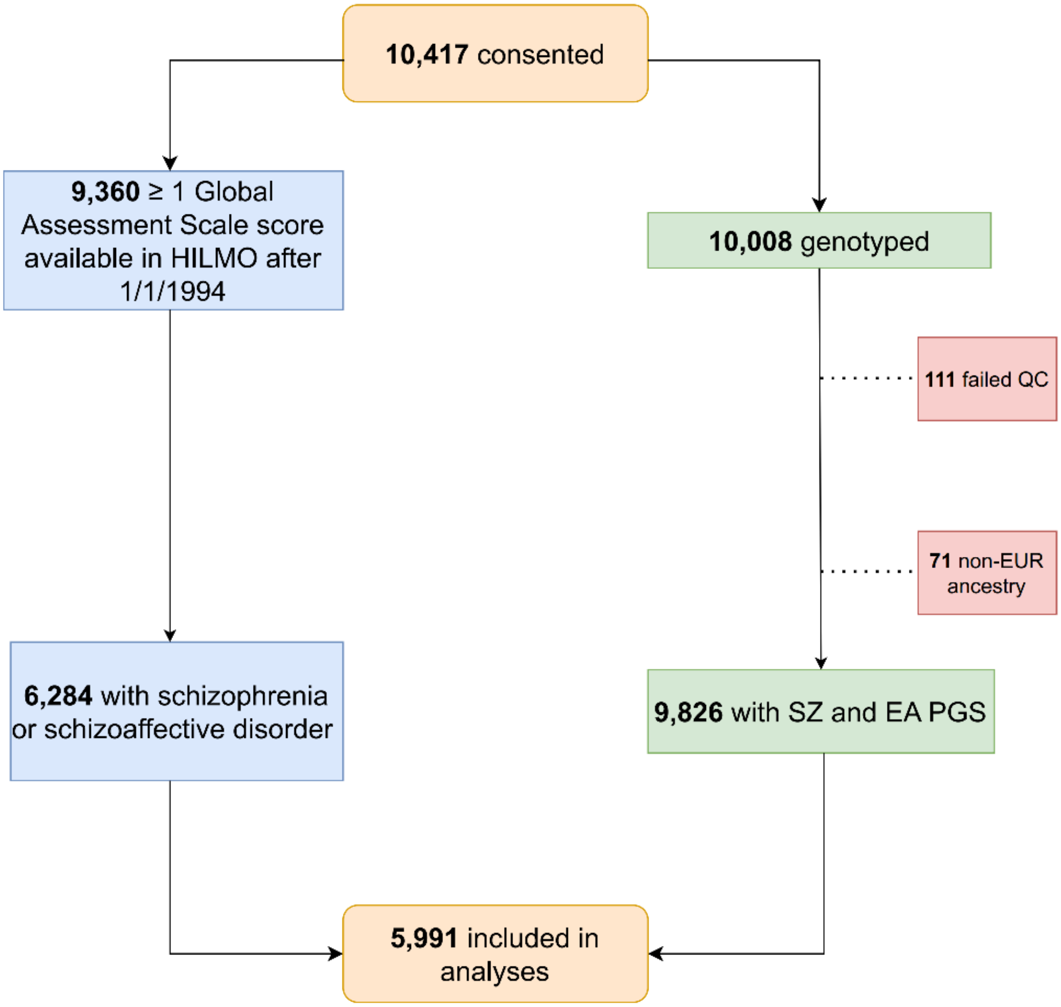


HILMO: Care Register for Health Care. SZ: schizophrenia. EA: educational attainment. PGS: polygenic score. EUR: European. QC: quality control.

**eFigure 2. PGS Associations Stratified by Admission GF Tertile**

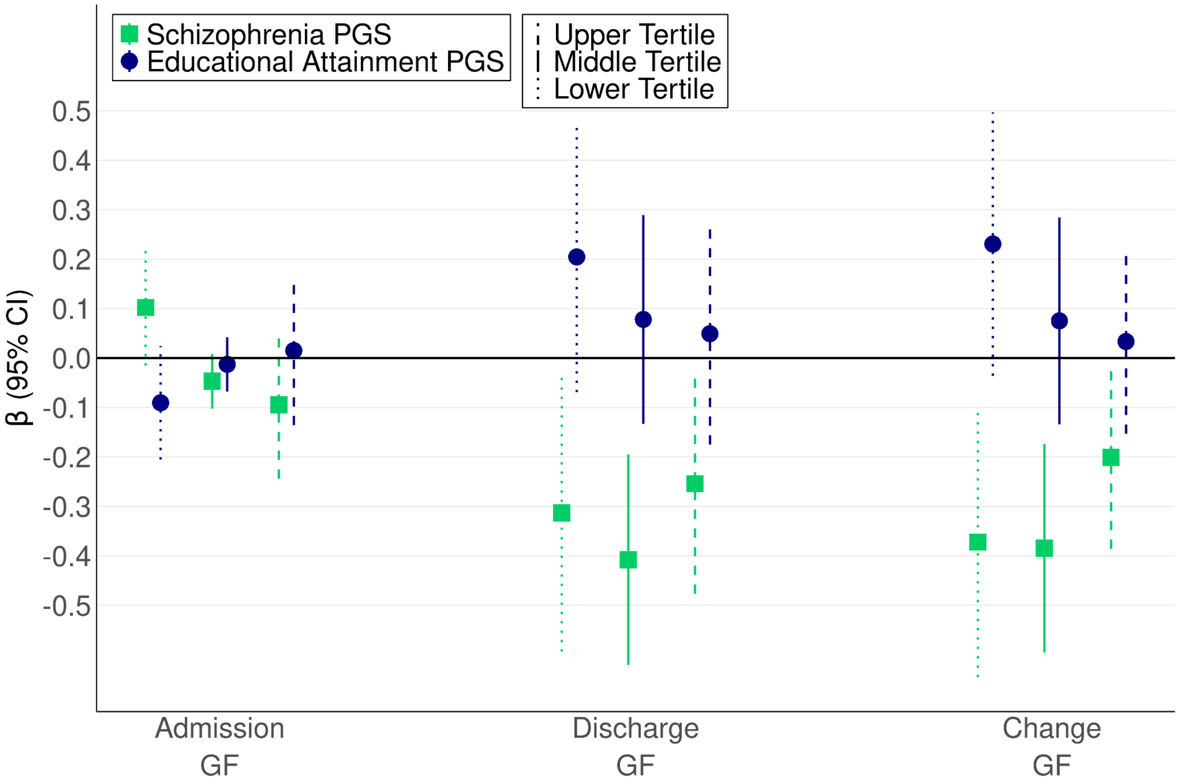


PGS: polygenic score. GF: global functioning. Change GF: Discharge GF adjusted for Admission GF. Bars depict 95% CIs.

#### eTables

##### eTable 1. Demographic and Descriptive Information

| Domain | Phenotype | n (%) | Median | IQR |
|---|---|---|---|---|
| Demographics | Sex (Female/Male) | 2733 (45.62) / 3258 (54.38) | - | - |
| | Age (Years) | 5991 (100) | 47 | 20 |
| | Diagnosis (Schizophrenia / Schizoaffective) | 5155 (86.05) / 836 (13.95) | - | - |
| Clinical Status and Inpatient Psychiatric History | Hospitalizations with GF (Post-December 31, 1993) | 5991 (100) | 6 | 9 |
| | Admission GF* | 5947 (99.27) | 39 | 16 |
| | Discharge GF* | 5991 (100) | 50 | 16 |
| | Length of Psychiatric Stay (Days)* | 5991 (100) | 14 | 38 |
| | Psychiatric Admissions First 5 years | 5991 (100) | 3 | 4 |
| | Days in Psychiatric Hospital First 5 years | 5991(100) | 158 | 290 |
| | Suicide Attempt History (Yes / No) | 3390 (56.58) / 2435 (40.64) | - | - |
| Education | Educational Attainment (Basic / Secondary / Higher) | 2238 (37.36) / 2681 (44.75) / 1018 (16.99) | - | - |
| | School Difficulties History (Yes / No) | 2837 (47.35) / 2990 (49.91) | - | - |
| CANTAB | RTI Median Reaction Time (ms) | 3921 (65.45) | 436 | 88 |
| | RTI All Errors | 3921 (65.45) | 0 | 1 |
| | PAL Patterns Reached | 3732 (62.29) | 6 | 4 |
| | PAL Total Attempts | 3732 (62.29) | 7 | 3 |
| | PAL First Attempt Memory | 3732 (62.29) | 7 | 7 |
| | PAL Total Errors Adjusted | 3732 (62.29) | 40 | 37 |

IQR: interquartile range. GF: global functioning. CANTAB: Cambridge Neuropsychological Test Automated Battery. RTI: Reaction Time. PAL: Paired Associates Learning. * To account for repeated measures when estimating descriptive statistics for these variables, we performed cluster bootstrapping by resampling at the individual participant level (1000 iterations).[19]

#### eTable 2. Estimates from Admission GF Linear Mixed-Effects Model

| Fixed Effects | | | | | |
|---|---|---|---|---|---|
| | Estimate/β | SE | 95% CI | *t* | *P* |
| Intercept | 163.62 | 14.38 | 135.43 to 191.81 | 11.38 | < .001 |
| SZ PGS | −0.20 | 0.10 | −0.40 to $-7.28 \times 10^{-6}$ | −1.96 | .050 |
| EA PGS | −0.23 | 0.10 | −0.43 to −0.04 | −2.32 | .020 |
| Sex | −0.13 | 0.20 | −0.52 to 0.26 | −0.67 | .502 |
| Year of Birth | −0.06 | 0.01 | −0.08 to −0.05 | −8.82 | < .001 |
| PC 1 | 62.93 | 14.31 | 34.87 to 91.00 | 4.40 | < .001 |
| PC 2 | −115.28 | 13.22 | −141.21 to −89.36 | −8.72 | < .001 |
| PC 3 | 108.66 | 13.34 | 82.50 to 134.81 | 8.14 | < .001 |
| PC 4 | −4.03 | 14.53 | −32.51 to 24.45 | −0.28 | .782 |
| PC 5 | −56.03 | 13.18 | −81.87 to −30.19 | −4.25 | < .001 |
| PC 6 | 27.86 | 13.17 | 2.04 to 53.68 | 2.12 | .035 |
| PC 7 | 73.40 | 13.99 | 45.98 to 100.83 | 5.25 | < .001 |
| PC 8 | −25.88 | 13.37 | −52.10 to 0.33 | −1.94 | .053 |
| PC 9 | 8.71 | 10.06 | −11.01 to 28.43 | 0.87 | .039 |
| PC 10 | 67.94 | 12.67 | 43.08 to 92.80 | 5.36 | < .001 |
| Random Effect | | | | | |
| | | | | Variance | SD |
| Participant (Intercept) | | | | 36.12 | 6.01 |
| Goodness of Fit | | | | | |
| | | | | Marginal | Conditional |
| Pseudo-$R^2$ | | | | 0.02 | 0.27 |

SZ: schizophrenia. EA: educational attainment. PGS: polygenic score. PC: principal component. Sex was coded as a binary variable (Female/Male).

#### eTable 3. Estimates from Discharge GF Linear Mixed-Effects Model

| Fixed Effects | | | | | |
|---|---|---|---|---|---|
| | Estimate/β | SE | 95% CI | *t* | *P* |
| Intercept | 1.78 | 14.26 | −26.16 to 29.73 | 0.13 | .901 |
| SZ PGS | −0.36 | 0.10 | −0.56 to −0.17 | −3.60 | < .001 |
| EA PGS | 0.15 | 0.10 | −0.04 to 0.35 | 1.56 | .119 |
| Sex | −0.67 | 0.20 | −1.05 to -0.28 | −3.37 | < .001 |
| Year of Birth | 0.03 | 0.01 | 0.01 to 0.04 | 3.51 | < .001 |
| Length of Stay | −0.13 | 0.03 | −0.20 to −0.07 | −3.98 | < .001 |
| PC 1 | 98.10 | 14.15 | 70.36 to 125.85 | 6.93 | < .001 |
| PC 2 | −74.00 | 13.14 | −99.76 to −48.23 | −5.63 | < .001 |
| PC 3 | 35.98 | 13.25 | 10.01 to 61.95 | 2.72 | .007 |
| PC 4 | 31.60 | 14.46 | 3.25 to 59.94 | 2.19 | .029 |
| PC 5 | −46.74 | 13.07 | −72.37 to −21.12 | −3.58 | < .001 |
| PC 6 | 24.62 | 13.08 | −1.03 to 50.26 | 1.88 | .060 |
| PC 7 | 62.32 | 13.85 | 35.16 to 89.48 | 4.50 | < .001 |
| PC 8 | 9.93 | 13.22 | −15.99 to 35.84 | 0.75 | .453 |
| PC 9 | 16.31 | 9.97 | −3.23 to 35.86 | 1.64 | .102 |
| PC 10 | 80.99 | 12.59 | 56.31 to 105.68 | 6.43 | < .001 |
| Random Effect | | | | | |
| | | | | Variance | SD |
| Participant (Intercept) | | | | 35.70 | 5.97 |
| Goodness of Fit | | | | | |
| | | | | Marginal | Conditional |
| Pseudo-$R^2$ | | | | 0.02 | 0.26 |

SZ: schizophrenia. EA: educational attainment. PGS: polygenic score. PC: principal component. Sex was coded as a binary variable (Female/Male). Length of stay was scaled in days and log-transformed.

#### eTable 4. Estimates from Change GF Linear Mixed-Effects Model

| Fixed Effects | | | | | |
|---|---|---|---|---|---|
| | Estimate/β | SE | 95% CI | *t* | *P* |
| Intercept | −63.44 | 12.49 | −87.93 to −38.95 | −5.08 | < .001 |
| SZ PGS | −0.31 | 0.09 | −0.49 to −0.14 | −3.56 | < .001 |
| EA PGS | 0.20 | 0.09 | 0.03 to 0.37 | 2.27 | .023 |
| Admission GF | 0.48 | 0.004 | 0.47 to 0.49 | 129.18 | < .001 |
| Sex | −0.62 | 0.17 | −0.96 to −0.28 | −3.60 | < .001 |
| Year of Birth | 0.05 | 0.006 | 0.04 to 0.06 | 7.52 | < .001 |
| Length of Stay | 1.05 | 0.03 | 0.99 to 1.12 | 33.16 | < .001 |
| PC 1 | 68.28 | 12.43 | 43.91 to 92.65 | 5.49 | < .001 |
| PC 2 | −24.57 | 11.49 | −47.09 to −2.05 | −2.14 | .033 |
| PC 3 | −12.52 | 11.59 | −35.24 to 10.20 | −1.08 | .280 |
| PC 4 | 28.03 | 12.61 | 3.30 to 52.75 | 2.22 | .026 |
| PC 5 | −22.77 | 11.44 | −45.21 to −0.34 | −1.99 | .047 |
| PC 6 | 9.57 | 11.44 | −12.84 to 31.99 | 0.84 | .403 |
| PC 7 | 28.68 | 12.15 | 4.86 to 52.50 | 2.36 | .018 |
| PC 8 | 19.87 | 11.60 | −2.88 to 42.62 | 1.71 | .087 |
| PC 9 | 10.81 | 8.73 | −6.31 to 27.93 | 1.24 | .216 |
| PC 10 | 50.02 | 11.01 | 28.43 to 71.62 | 4.54 | < .001 |
| Random Effect | | | | | |
| | | | | Variance | SD |
| Participant (Intercept) | | | | 27.65 | 5.26 |
| Goodness of Fit | | | | | |
| | | | | Marginal | Conditional |
| Pseudo-$R^2$ | | | | 0.24 | 0.43 |

Change GF: Discharge GF adjusted for Admission GF. SZ: schizophrenia. EA: educational attainment. PGS: polygenic score. PC: principal component. Sex was coded as a binary variable (Female/Male). Length of stay was scaled in days and log-transformed.

#### eTable 5. Multiple Comparisons Correction

| Fixed Effects | | | |
|---|---|---|---|
| Predictor | Outcome | Unadjusted *P* | FDR-Adjusted *P* |
| SZ PGS | Admission GF | .0500 | .0600 |
| | Discharge GF* | .0003 | .0012 |
| | Change GF* | .0004 | .0012 |
| EA PGS | Admission GF* | .0204 | .0348 |
| | Discharge GF | .1191 | .1191 |
| | Change GF* | .0232 | .0348 |

SZ: schizophrenia. EA: educational attainment. PGS: polygenic score. GF: global functioning. Change GF: discharge GF adjusted for admission GF. FDR: false discovery rate, which we controlled using the Benjamini-Hochberg procedure.[20] *Significant at .05 after FDR adjustment.

**eTable 6. Estimates from Tertile-Stratified Admission GF Linear Mixed-Effects Model**

| Fixed Effects | | | | | | |
|---|---|---|---|---|---|---|
| | Tertile | Estimate/β | SE | 95% CI | *t* | *P* |
| Intercept | Upper | 107.74 | 11.63 | 84.95 to 130.53 | 9.27 | < .001 |
| | Middle | 45.03 | 4.13 | 36.93 to 53.12 | 10.90 | < .001 |
| | Lower | 45.97 | 8.44 | 29.42 to 62.51 | 5.45 | < .001 |
| SZ PGS | Upper | 0.09 | 0.08 | −0.24 to 0.05 | −1.24 | .214 |
| | Middle | −0.05 | 0.03 | −0.10 to 0.01 | -0.45 | .097 |
| | Lower | 0.10 | 0.06 | −0.02 to 0.22 | 1.70 | .088 |
| EA PGS | Upper | 0.02 | 0.08 | −0.14 to 0.17 | 0.20 | .845 |
| | Middle | −0.01 | 0.03 | −0.07 to 0.04 | −0.45 | .650 |
| | Lower | −0.09 | 0.06 | −0.21 to 0.02 | −1.55 | .122 |
| Sex | Upper | 0.08 | 0.15 | −0.22 to 0.38 | 0.53 | .596 |
| | Middle | −0.10 | 0.06 | −0.20 to 0.01 | −1.72 | .085 |
| | Lower | −0.30 | 0.12 | −0.53 to −0.07 | −2.57 | .010 |
| Year of Birth | Upper | −0.03 | 0.01 | −0.04 to −0.02 | −4.82 | < .001 |
| | Middle | −0.003 | 0.002 | −0.01 to 0.001 | −1.66 | .098 |
| | Lower | −0.01 | 0.004 | −0.02 to −0.002 | −2.45 | .015 |
| PC 1 | Upper | 32.36 | 11.11 | 10.58 to 54.14 | 2.91 | .004 |
| | Middle | 14.29 | 4.05 | 6.34 to 22.24 | 3.52 | < .001 |
| | Lower | 18.42 | 8.40 | 1.95 to 34.90 | 2.19 | .028 |
| PC 2 | Upper | 0.002 | 10.23 | −20.05 to 20.06 | $1.65 \times 10^{-4}$ | 1.00 |
| | Middle | −1.18 | 3.81 | −8.65 to 6.30 | −0.31 | .758 |
| | Lower | −22.12 | 7.91 | −37.63 to −6.61 | −2.80 | .005 |
| PC 3 | Upper | 13.50 | 10.69 | −7.46 to 34.46 | 1.26 | .207 |
| | Middle | 19.85 | 3.87 | 12.25 to 27.44 | 5.12 | < .001 |
| | Lower | 6.15 | 7.80 | −9.14 to 21.44 | 0.79 | .431 |
| PC 4 | Upper | 21.76 | 10.88 | 0.43 to 43.10 | 2.00 | .046 |
| | Middle | −8.50 | 4.35 | −17.03 to 0.02 | −1.96 | .051 |

| | Tertile | Estimate/β | SE | 95% CI | *t* | *P* |
|---|---|---|---|---|---|---|
| | Lower | 8.38 | 8.84 | −8.95 to 25.72 | 0.95 | .343 |
| PC 5 | Upper | 1.23 | 10.08 | −18.53 to 20.98 | 0.12 | .903 |
| | Middle | 4.16 | 3.74 | −3.17 to 11.49 | 1.11 | .266 |
| | Lower | 0.53 | 7.85 | −14.87 to 15.93 | 0.07 | .946 |
| PC 6 | Upper | 21.64 | 9.58 | 2.86 to 40.42 | 2.26 | .024 |
| | Middle | 1.73 | 3.82 | −5.75 to 9.21 | 0.45 | .650 |
| | Lower | 15.96 | 8.07 | 0.14 to 31.78 | 1.98 | .048 |
| PC 7 | Upper | −6.64 | 10.67 | −27.56 to 14.27 | −0.62 | .534 |
| | Middle | 8.94 | 3.94 | 1.22 to 16.67 | 2.27 | .023 |
| | Lower | 27.06 | 8.29 | 10.82 to 43.31 | 3.27 | .001 |
| PC 8 | Upper | 9.46 | 10.53 | −11.18 to 30.10 | 0.90 | .369 |
| | Middle | −4.69 | 3.77 | −12.09 to 2.70 | −1.24 | .214 |
| | Lower | 7.35 | 7.89 | −8.12 to 22.82 | 0.93 | .352 |
| PC 9 | Upper | −9.29 | 8.09 | −25.16 to 6.58 | −1.15 | .251 |
| | Middle | 3.20 | 2.90 | −2.48 to 8.88 | 1.10 | .270 |
| | Lower | 3.73 | 5.39 | −6.84 to 14.29 | 0.69 | .489 |
| PC 10 | Upper | 5.51 | 9.12 | −12.38 to 23.39 | 0.60 | .546 |
| | Middle | 4.30 | 3.54 | −2.65 to 11.25 | 1.21 | .225 |
| | Lower | 16.13 | 7.75 | 0.94 to 31.31 | 2.08 | .037 |
| Random Effect | | | | | | |
| | | | | | Variance | SD |
| Participant (Intercept) | Upper | | | | 7.96 | 2.82 |
| | Middle | | | | 0.93 | 0.96 |
| | Lower | | | | 5.18 | 2.28 |
| Goodness of Fit | | | | | | |
| | | | | | Marginal | Conditional |
| Pseudo-$R^2$ | Upper | | | | 0.01 | 0.19 |

| | | | Marginal | Conditional |
|---|---|---|---|---|
| | Middle | | 0.01 | 0.11 |
| | Lower | | 0.004 | 0.13 |

SZ: schizophrenia. EA: educational attainment. PGS: polygenic score. PC: principal component. Sex was coded as a binary variable (Female/Male).

#### eTable 7. Estimates from Tertile-Stratified Discharge GF Linear Mixed-Effects Model

| Fixed Effects | | | | | | |
|---|---|---|---|---|---|---|
| | Tertile | Estimate/β | SE | 95% CI | *t* | *P* |
| Intercept | Upper | 48.02 | 17.11 | 14.48 to 81.55 | 2.81 | .005 |
| | Middle | −26.02 | 15.63 | −56.66 to 4.61 | −1.66 | .096 |
| | Lower | −77.29 | 20.02 | −116.53 to −38.05 | −3.86 | < .001 |
| SZ PGS | Upper | −0.25 | 0.11 | −0.48 to −0.03 | −2.24 | .025 |
| | Middle | −0.41 | 0.11 | −0.62 to −0.19 | −3.75 | < .001 |
| | Lower | −0.31 | 0.14 | −0.59 to −0.03 | −2.19 | .028 |
| EA PGS | Upper | 0.05 | 0.11 | −0.18 to 0.27 | 0.43 | .667 |
| | Middle | 0.08 | 0.11 | −0.13 to 0.29 | 0.72 | .469 |
| | Lower | 0.20 | 0.14 | −0.07 to 0.48 | 1.46 | .143 |
| Sex | Upper | −0.37 | 0.23 | −0.81 to 0.07 | −1.63 | .103 |
| | Middle | −0.63 | 0.21 | −1.04 to −0.21 | −2.94 | .003 |
| | Lower | −0.86 | 0.28 | −1.40 to −0.31 | −3.08 | .002 |
| Year of Birth | Upper | 0.00 | 0.01 | −0.01 to 0.02 | 0.54 | .586 |
| | Middle | 0.04 | 0.01 | 0.02 to 0.05 | 4.73 | < .001 |
| | Lower | 0.06 | 0.01 | 0.04 to 0.08 | 6.06 | < .001 |
| Length of Stay | Upper | 0.21 | 0.05 | 0.11 to 0.31 | 4.10 | < .001 |
| | Middle | 0.99 | 0.05 | 0.89 to 1.09 | 19.82 | < .001 |
| | Lower | 0.89 | 0.07 | 0.76 to 1.02 | 13.33 | < .001 |
| PC 1 | Upper | 65.98 | 16.48 | 33.67 to 98.30 | 4.00 | < .001 |
| | Middle | 58.94 | 15.48 | 28.60 to 89.29 | 3.81 | < .001 |
| | Lower | 95.05 | 20.00 | 55.84 to 134.27 | 4.75 | < .001 |
| PC 2 | Upper | 4.61 | 15.05 | −24.89 to 34.12 | 0.31 | .759 |
| | Middle | −15.89 | 14.49 | −44.30 to 12.51 | −1.10 | .273 |
| | Lower | −55.31 | 18.74 | −92.05 to −18.57 | −2.95 | .003 |
| PC 3 | Upper | 13.70 | 15.65 | −16.98 to 44.39 | 0.88 | .381 |

| | Tertile | Estimate/β | SE | 95% CI | *t* | *P* |
|---|---|---|---|---|---|---|
| | Middle | −6.10 | 14.75 | −35.02 to 22.82 | −0.41 | .679 |
| | Lower | −18.63 | 18.52 | −54.93 to 17.67 | −1.01 | .314 |
| PC 4 | Upper | 73.95 | 16.11 | 42.37 to 105.53 | 4.59 | < .001 |
| | Middle | 18.36 | 16.31 | −13.61 to 50.33 | 1.13 | .260 |
| | Lower | 14.54 | 20.77 | −26.18 to 55.27 | 0.70 | .484 |
| PC 5 | Upper | 6.68 | 14.89 | −22.52 to 35.88 | 0.45 | .654 |
| | Middle | −20.29 | 14.32 | −48.37 to 7.79 | −1.42 | .157 |
| | Lower | −32.34 | 18.64 | −68.89 to 4.21 | −1.73 | .083 |
| PC 6 | Upper | 23.20 | 14.19 | −4.63 to 51.02 | 1.63 | .102 |
| | Middle | −4.57 | 14.52 | −33.02 to 23.89 | −0.31 | .753 |
| | Lower | 32.86 | 19.07 | −4.52 to 70.24 | 1.72 | .085 |
| PC 7 | Upper | −11.00 | 15.84 | −42.06 to 20.06 | −0.69 | .488 |
| | Middle | 38.67 | 15.15 | 8.96 to 68.38 | 2.55 | .011 |
| | Lower | 60.20 | 19.69 | 21.60 to 98.79 | 3.06 | .002 |
| PC 8 | Upper | 11.42 | 15.55 | −19.08 to 41.92 | 0.73 | .463 |
| | Middle | 4.98 | 14.40 | -23.25 to 33.21 | 0.35 | .730 |
| | Lower | 37.09 | 18.77 | 0.29 to 73.88 | 1.98 | .048 |
| PC 9 | Upper | −7.00 | 11.78 | −30.10 to 16.10 | −0.59 | .553 |
| | Middle | 22.72 | 11.19 | 0.77 to 44.67 | 2.03 | .043 |
| | Lower | 16.20 | 13.00 | −9.28 to 41.69 | 1.25 | .213 |
| PC 10 | Upper | 22.93 | 13.60 | −3.72 to 49.59 | 1.69 | .092 |
| | Middle | 63.89 | 13.56 | 37.31 to 90.46 | 4.71 | < .001 |
| | Lower | 65.73 | 18.34 | 29.77 to 101.68 | 3.58 | < .001 |
| Random Effect | | | | | | |
| | | | | | Variance | SD |
| Participant (Intercept) | Upper | | | | 23.44 | 4.84 |
| | Middle | | | | 26.48 | 5.15 |

| | | | Variance | SD |
|---|---|---|---|---|
| | Lower | | 46.43 | 6.81 |
| Goodness of Fit | | | | |
| | | | Marginal | Conditional |
| Pseudo-$R^2$ | Upper | | 0.01 | 0.30 |
| | Middle | | 0.03 | 0.31 |
| | Lower | | 0.02 | 0.28 |

SZ: schizophrenia. EA: educational attainment. PGS: polygenic score. PC: principal component. Sex was coded as a binary variable (Female/Male). Length of stay was scaled in days and log-transformed.

#### eTable 8. Estimates from Tertile-Stratified Change GF Linear Mixed-Effects Model

| Fixed Effects | | | | | | |
|---|---|---|---|---|---|---|
| | Tertile | Estimate/β | SE | 95% CI | *t* | *P* |
| Intercept | Upper | -25.58 | 14.24 | -53.49 to 2.33 | -1.80 | .073 |
| | Middle | -47.47 | 15.51 | -77.87 to -17.07 | -3.06 | .002 |
| | Lower | -97.62 | 19.50 | -135.84 to -59.41 | -5.01 | < .001 |
| SZ PGS | Upper | -0.20 | 0.09 | -0.39 to -0.02 | -2.12 | .034 |
| | Middle | -0.38 | 0.11 | -0.60 to -0.17 | -3.57 | < .001 |
| | Lower | -0.37 | 0.14 | -0.65 to -0.10 | -2.68 | .008 |
| EA PGS | Upper | 0.03 | 0.10 | -0.15 to 0.22 | 0.35 | .726 |
| | Middle | 0.08 | 0.11 | -0.13 to 0.28 | 0.70 | .481 |
| | Lower | 0.23 | 0.14 | -0.04 to 0.50 | 1.70 | .090 |
| Admission GF | Upper | 0.73 | 0.01 | 0.71 to 0.74 | 95.33 | < .001 |
| | Middle | 0.48 | 0.02 | 0.44 to 0.52 | 22.70 | < .001 |
| | Lower | 0.44 | 0.01 | 0.41 to 0.47 | 31.36 | < .001 |
| Sex | Upper | -0.42 | 0.19 | -0.79 to -0.05 | -2.24 | .025 |
| | Middle | -0.57 | 0.21 | -0.98 to -0.16 | -2.70 | .007 |
| | Lower | -0.72 | 0.27 | -1.25 to -0.18 | -2.64 | .008 |
| Year of Birth | Upper | 0.02 | 0.01 | 0.01 to 0.04 | 3.09 | .002 |
| | Middle | 0.04 | 0.01 | 0.02 to 0.05 | 4.96 | < .001 |
| | Lower | 0.07 | 0.01 | 0.05 to 0.09 | 6.66 | < .001 |
| Length of Stay | Upper | 0.77 | 0.04 | 0.69 to 0.86 | 18.10 | < .001 |
| | Middle | 1.11 | 0.05 | 1.02 to 1.21 | 22.47 | < .001 |
| | Lower | 1.08 | 0.07 | 0.96 to 1.21 | 16.63 | < .001 |
| PC 1 | Upper | 44.08 | 13.71 | 17.19 to 70.97 | 3.21 | .001 |
| | Middle | 51.63 | 15.33 | 21.57 to 81.69 | 3.37 | < .001 |
| | Lower | 86.04 | 19.47 | 47.87 to 124.21 | 4.42 | < .001 |
| PC 2 | Upper | 5.50 | 12.52 | -19.05 to 30.05 | 0.44 | .661 |
| | Middle | -14.22 | 14.35 | -42.36 to 13.92 | -0.99 | .322 |

| | Tertile | Estimate/β | SE | 95% CI | *t* | *P* |
|---|---|---|---|---|---|---|
| | Lower | -46.87 | 18.24 | -82.63 to -11.11 | -2.57 | .010 |
| PC 3 | Upper | 2.91 | 13.01 | -22.61 to 28.42 | 0.22 | .823 |
| | Middle | -15.41 | 14.62 | -44.08 to 13.25 | -1.05 | .292 |
| | Lower | -21.25 | 18.02 | -56.58 to 14.07 | -1.18 | .238 |
| PC 4 | Upper | 55.74 | 13.42 | 29.43 to 82.05 | 4.15 | < .001 |
| | Middle | 21.77 | 16.15 | -9.89 to 53.43 | 1.35 | .178 |
| | Lower | 9.59 | 20.21 | -30.04 to 49.22 | 0.47 | .635 |
| PC 5 | Upper | 6.90 | 12.39 | -17.40 to 31.20 | 0.56 | .578 |
| | Middle | -21.30 | 14.19 | -49.12 to 6.52 | -1.50 | .133 |
| | Lower | -31.94 | 18.15 | -67.52 to 3.63 | -1.76 | .078 |
| PC 6 | Upper | 8.28 | 11.82 | -14.90 to 31.46 | 0.70 | .484 |
| | Middle | -6.10 | 14.38 | -34.29 to 22.09 | -0.42 | .671 |
| | Lower | 25.03 | 18.56 | -11.35 to 61.41 | 1.35 | .178 |
| PC 7 | Upper | -7.14 | 13.19 | -33.00 to 18.72 | -0.54 | .588 |
| | Middle | 34.58 | 15.01 | 5.14 to 64.01 | 2.30 | .021 |
| | Lower | 48.93 | 19.16 | 11.35 to 86.50 | 2.55 | .011 |
| PC 8 | Upper | 4.28 | 12.94 | -21.09 to 29.65 | 0.33 | .741 |
| | Middle | 7.10 | 14.26 | -20.87 to 35.06 | 0.50 | .619 |
| | Lower | 33.88 | 18.27 | -1.93 to 69.69 | 1.85 | .064 |
| PC 9 | Upper | -0.59 | 9.80 | -19.79 to 18.62 | -0.06 | .952 |
| | Middle | 21.47 | 11.09 | -0.28 to 43.22 | 1.94 | .053 |
| | Lower | 14.52 | 12.65 | -10.29 to 39.32 | 1.15 | .251 |
| PC 10 | Upper | 18.71 | 11.33 | -3.51 to 40.92 | 1.65 | .099 |
| | Middle | 62.15 | 13.43 | 35.83 to 88.48 | 4.63 | < .001 |
| | Lower | 58.15 | 17.85 | 23.16 to 93.14 | 3.26 | .001 |

| Random Effect | | | | |
|---|---|---|---|---|
| | | | Variance | SD |
| Participant (Intercept) | Upper | | 17.31 | 4.16 |
| | Middle | | 26.27 | 5.13 |
| | Lower | | 44.00 | 6.63 |
| Goodness of Fit | | | | |
| | | | Marginal | Conditional |
| Pseudo-$R^2$ | Upper | | 0.30 | 0.53 |
| | Middle | | 0.05 | 0.33 |
| | Lower | | 0.07 | 0.31 |

Change GF: Discharge GF adjusted for Admission GF. SZ: schizophrenia. EA: educational attainment. PGS: polygenic score. PC: principal component. Sex was coded as a binary variable (Female/Male). Length of stay was scaled in days and log-transformed.
