## Supplementary material for "Polygenic Scores for Schizophrenia and Educational Attainment Predict Global Functioning Across Psychiatric Hospitalization Among People with Schizophrenia": Figures 1-3

Figure 1. Global Functioning Across 59,795 Psychiatric Hospitalizations

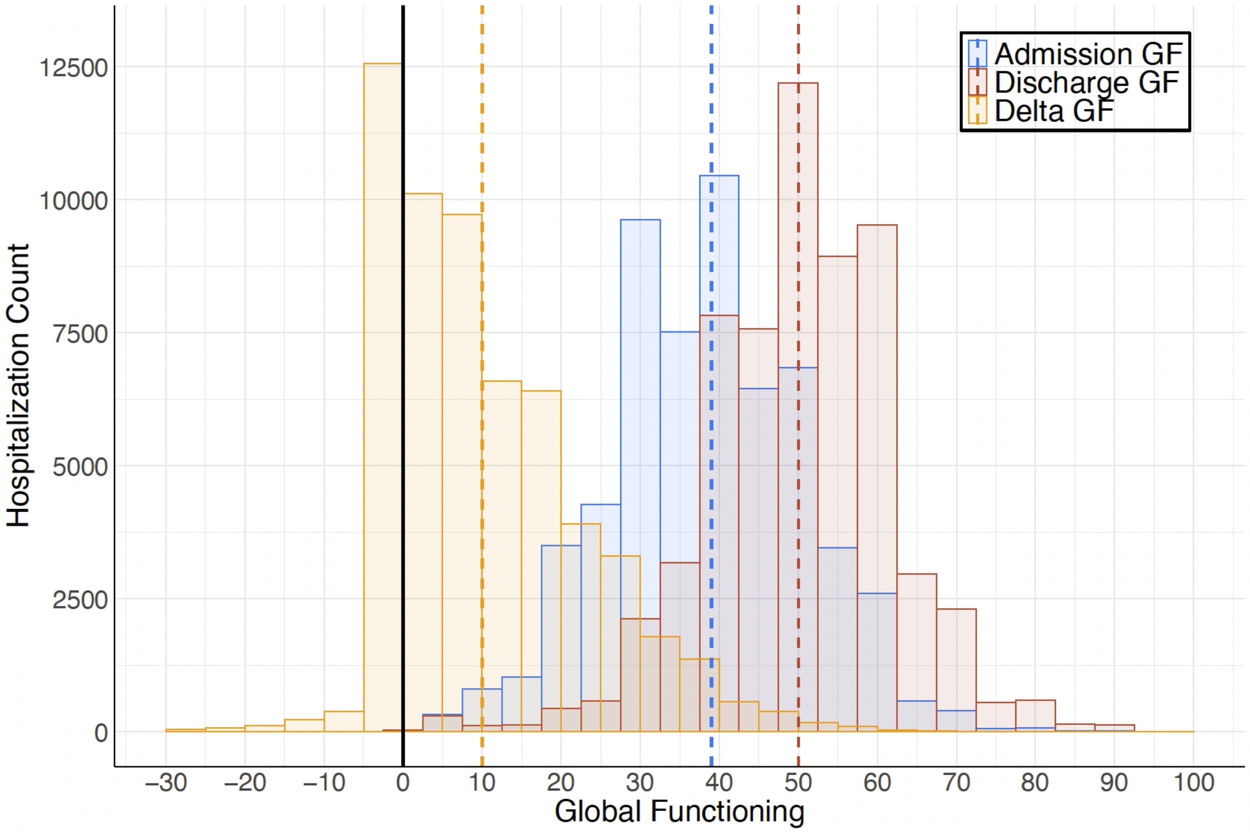


GF: global functioning. Delta GF = Discharge GF - Admission GF. Dashed vertical lines depict median GF scores.

Figure 2. Phenotypic Associations Among Global Functioning Scores and Deep Phenotypes

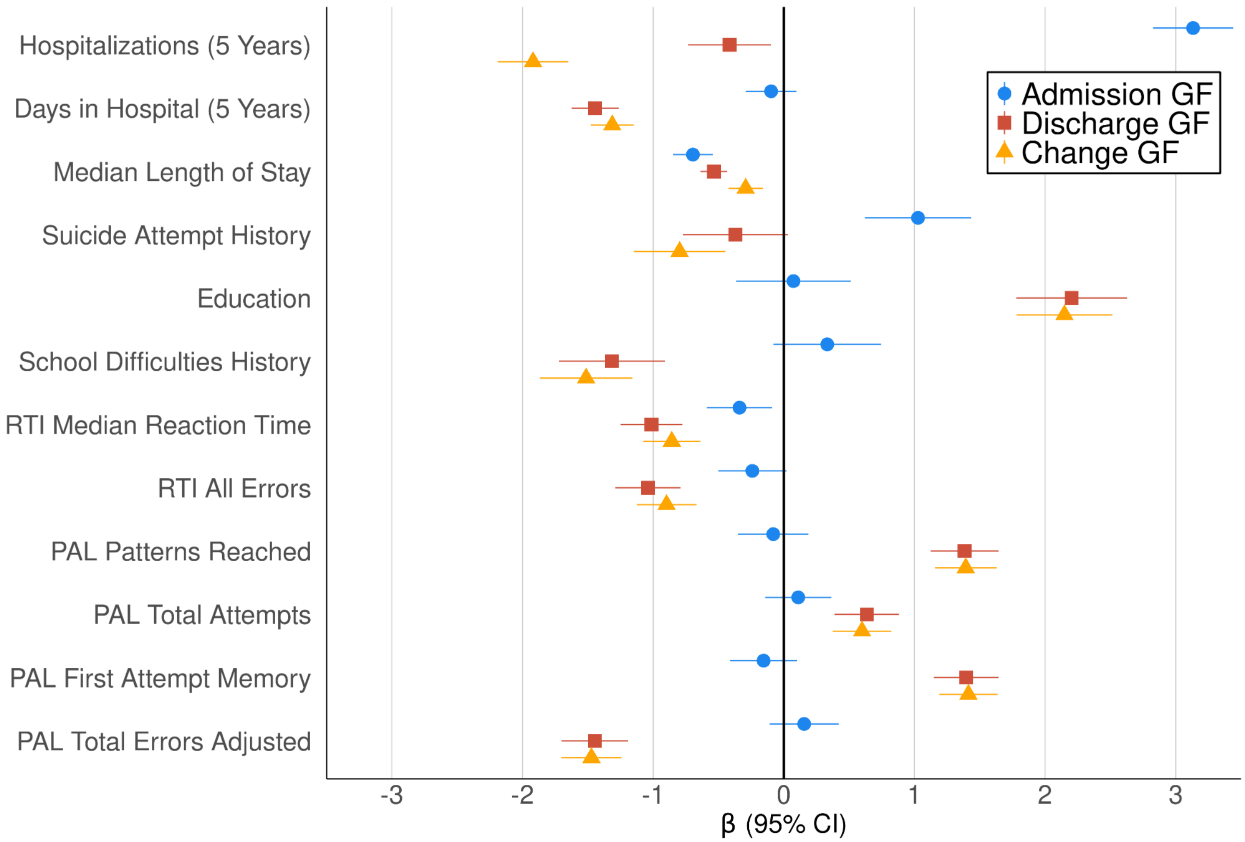


GF: global functioning. Change GF = Discharge GF adjusted for Admission GF. RTI: Reaction Time. PAL: Paired Associates Learning. 5 Years: these variables were calculated for psychiatric hospitalizations within the initial 5 years following a participant's first admission (Supplement). RTI and PAL were assessed at study entry (between November 2015 and December 2018). Bars depict 95% CIs

Figure 3. Schizophrenia and EA PGS Predict Global Functioning Across Hospitalization

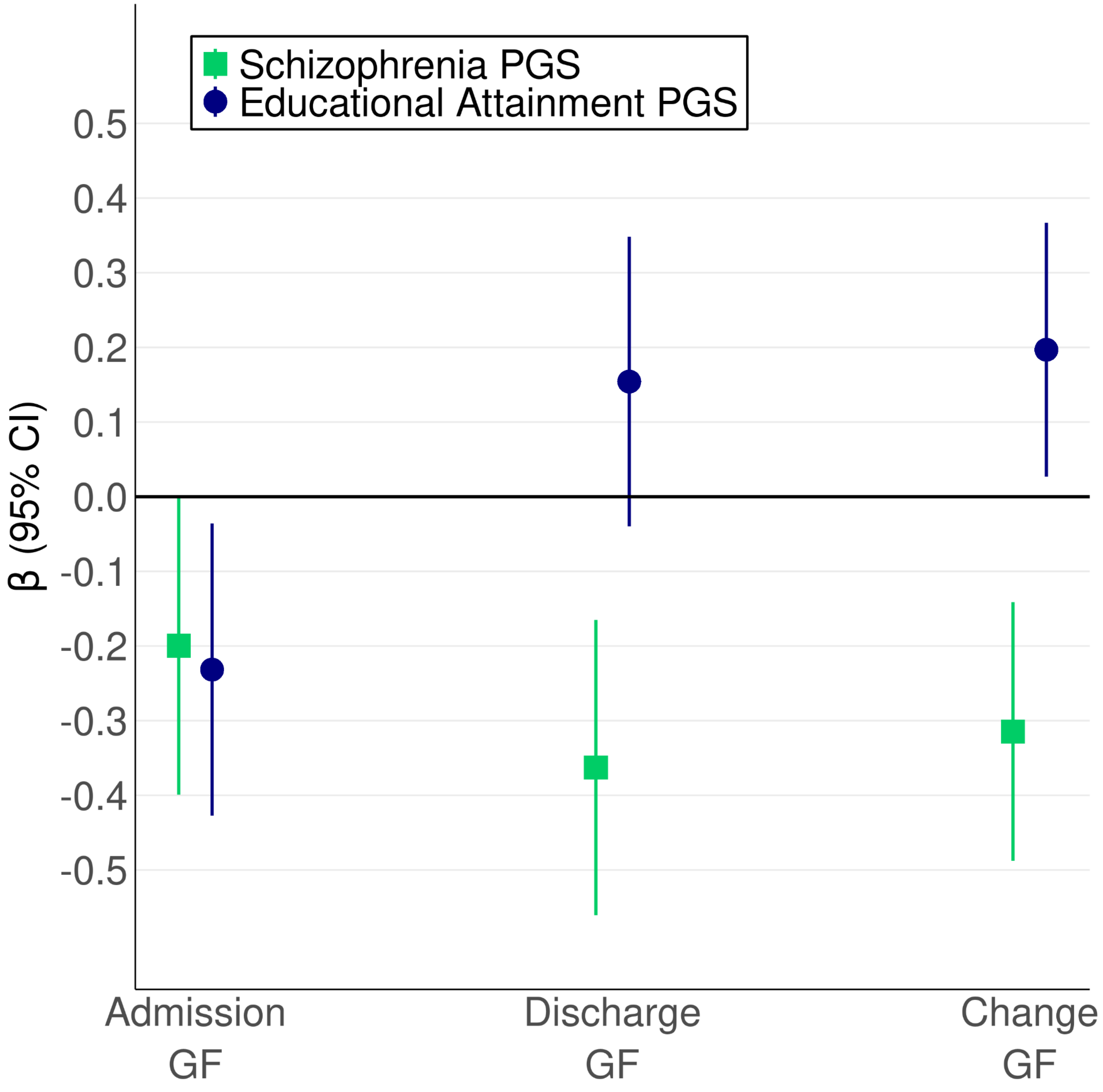


PGS: polygenic score. GF: global functioning. Change GF: Discharge GF adjusted for Admission GF. Bars depict 95% CIs.
